# Dopa responsiveness in Parkinson’s disease

**DOI:** 10.1101/2024.04.26.24306435

**Authors:** Sacha E Gandhi, Anahita Nodehi, Michael A Lawton, Katherine A Grosset, Vicky Marshall, Yoav Ben-Shlomo, Donald G Grosset

## Abstract

**Background:** Dopaminergic responsiveness is a defining feature of Parkinson’s disease (PD). However, there is limited information on how this evolves over time.

**Objectives:** To examine serial dopaminergic responses, if there are distinct patterns, and which factors predict these.

**Methods:** We analyzed data from the *Parkinson’s Progression Markers Initiative* on repeated dopaminergic challenge tests (≥ 24.5% defined as a definite response). Growth-mixture modeling evaluated for different response patterns and multinomial logistic regression tested for predictors of these clusters.

**Results:** There were 1,525 dopaminergic challenge tests from 336 patients. At enrolment, mean age was 61.2 years (SD 9.6), 66.4% were male and disease duration was 0.5 years (SD 0.5). 1 to 2 years after diagnosis, 48.0% of tests showed a definite response, but this proportion increased with longer duration (51.1-74.3%). We identified 3 response groups: ‘Striking’ (n = 29, 8.7%); ‘Excellent’ (n = 110; 32.7%) and ‘Modest’ (n = 197, 58.6%). Significant differences were as follows: striking responders commenced treatment earlier (P = 0.02), were less likely to be on dopamine agonist monotherapy (P = 0.01), and had better cognition (P < 0.01) and activities of daily living (P = 0.01). Excellent responders had higher challenge doses (P = 0.03) and were more likely to be on combination therapy (P < 0.01).

**Conclusion:** Three distinct patterns of the dopaminergic response were observed. The proportion of PD cases with definite dopa responsiveness increased over time, so the initial treatment response may be an unreliable diagnostic aid.

---

A good dopaminergic response typifies Parkinson’s disease and is a key part of the diagnostic process^1^. Variable responses are reported in clinical trials^2^, cohort^3^ and autopsy studies^4, 5^ and impact upon clinical outcomes^2, 3^. However, information is limited on the evolution of the dopaminergic response in PD. Some patients fail to respond to an acute L-dopa challenge in early PD but subsequently respond well to chronic dopaminergic therapy^6, 7^. Methods of assessing the dopaminergic response vary. An overall grading of the L-dopa response is reported in two studies with autopsy diagnostic confirmation, either in 3 (marked, moderate, and nil response)^5^ or 4 response groups (definite, good, limited and nil-to-poor)^4^, while other studies reported cross-sectional findings from on-off motor scoring^3, 8^. Previous observational studies that have used challenge tests to evaluate the response over time involved small cohorts and applied the superseded Modified Webster scale^9–13^. A previous report showed that the response amplitude increased over the first 5 years but then remained steady, although longer-term data was available in only 24 cases^11^.

The topic of longitudinal dopaminergic responsiveness is important for two main reasons. First, there are clinical implications: patients with a limited response in early disease have greater motor severity, faster motor progression^3^ and a lower likelihood of motor complications^11^. Second, L-dopa responsiveness is a key feature that helps to distinguish PD from other degenerative parkinsonian conditions where the response is more limited and wanes over time. The MDS diagnostic criteria define a clear and dramatic beneficial response to dopaminergic therapy as a supportive criterion for PD, and the absence of an observable response to high dose L-dopa despite at least moderate disease severity an absolute exclusion^1^. The finding, however, that a small subset of autopsy-proven PD cases appear not to respond to L-dopa^5^, or have a nil-to-poor response^4^, questions the concept of dopa- responsiveness being a defining feature of PD.

The *Parkinson’s Progression Marker Initiative* (*PPMI*) study has performed standardized serial on-off scoring in relation to prevailing dopaminergic therapy, providing unique prospective information about both initial and subsequent treatment responses^14^. We analyzed *PPMI* data with 3 aims: (1) to examine whether dopaminergic responsiveness changes over time; (2) to examine if there are, or are not, distinct patterns of the dopaminergic response, using a data-driven analytical approach; and (3) assuming different groups, to test for clinical characteristics as predictors of these treatment response groups.

## Methods

*PPMI* is an international multicenter observational study, in which 423 PD patients from 24 centers were followed longitudinally to establish clinical, biospecimen, imaging, and genetic biomarkers of progression^14, 15^. Patients were recruited up to 2 years after diagnosis if they were 30 years or older, untreated, and if they had at least two of bradykinesia, rest tremor or rigidity, or, alternatively, either asymmetric bradykinesia or an asymmetric rest tremor. Patients were required to have a dopaminergic deficit in dopamine transporter (DaT) imaging with ^123^I Ioflupane (or vesicular monoamine transporter (VMAT-2) imaging with ^18^F AV133 in Australia).

Imaging data were analyzed by the *PPMI* Imaging Core and defined against pre-specified criteria: visual assessment combined with uptake ratios (< 65% of age-expected putamen binding ratio). Exclusion criteria included a diagnosis of dementia or drug-induced parkinsonism. Patients were followed for up to 13 years, with assessments 3-monthly during the first year and 6-monthly thereafter.

Participants underwent comprehensive clinical assessment, including parts I to IV of the Movement Disorders Society-Unified Parkinson’s Disease Rating Scale (MDS UPDRS), with part III (motor) scores in the ‘on’ and ‘off’’ state^16^. Challenge tests were performed six-monthly to assess dopaminergic responsiveness over time.

Cognition was assessed with the Montreal Cognitive assessment (MoCA), adjusted for years of education. *PPMI* data were accessed in November 2022 at https://www.ppmi-info.org/^15^.

### Measured responses

The effect of dopaminergic therapy (both levodopa and dopamine agonists) was calculated using the following formula:

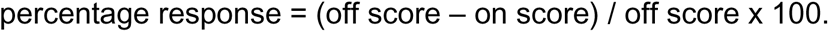

Patients withheld their dopaminergic therapy overnight for at least 6 hours (or 12 hours for long-acting drugs) prior to being scored in the ‘off’ state and were then scored in the ‘on’ state 1.5 hours after taking their usual morning dose (L-dopa, dopamine agonist, or both). For patients with deep brain stimulation (DBS), DBS was switched off 1 hour before ‘off’ scores were recorded and was switched on for assessment in the ‘on’ state. Alternatively, and less commonly, patients were scored in the ‘on’ state and waited until their medication wore off when an ‘off’ score was recorded. Levodopa equivalent doses (LED) were calculated by standard formulas^17^, updated for opicapone^18^.

Responses were analyzed against the 24.5% threshold in the MDS UPDRS part III score, which equates to 30% change in the UPDRS part III score^19^ and is defined in diagnostic criteria as representing a marked improvement^1^. Responses were declared ‘definite’ when the improvement in the MDS UPDRS part III score was ≥ 24.5% and ‘limited’ when < 24.5%, as reported previously^3^.

### Subjective grading

Clinicians were asked at repeated time points to answer yes, no, or not applicable to 2 questions relating to the treatment response: (a) ‘Is there little or no response to L-dopa?’ and, following a protocol update in June 2020, they were additionally asked (b) ‘Is there a clear and dramatic response to L-dopa?’. When there were consistent recordings of ‘little or no response’, a case was classed as having *nil response*. When there was ‘a clear and dramatic response’ at least once, the case was defined as responsive. The overall L-dopa response in individual cases was then categorised as *marked* when at least 50% of the subjective gradings recorded a ‘clear and dramatic response’, and *moderate* when this was less than 50%.

Subjective clinician assessments were compared with the measured responses from individual challenge tests when collected at the same visit. If a ‘clear and dramatic response’ was recorded as present, the test was graded *marked.* If ‘little or no response’ was recorded as present, the test was graded as showing *nil response.* If both ‘clear and dramatic response’ and ‘little or no response’ were recorded as ‘no’, the test was graded *moderate*. As the second question ‘Is there a clear and dramatic response to L-dopa?’ was introduced in June 2020, a grade of *moderate* could only be ascribed to cases evaluated after this date.

### Modeling approaches

A data-driven approach was used to identify if there were, or were not, multiple response clusters using only percentage response and disease duration in a growth mixture model (GMM)^20^. The model parameters were estimated using Maximum Likelihood Estimation (MLE) through the Expectation-Maximization (EM) algorithm, and the functional form of time was based on fractional polynomials^21^. In order to identify the number of classes that best fitted the data, we compared the following goodness-of-fit criteria: sample-size adjusted Bayesian Information Criterion (SABIC), entropy values, and the Akaike Information Criterion (AIC)^22, 23^.

After GMM was applied, univariable multinomial logistic regression was performed, using the largest cluster as the reference group, to evaluate the association between clinical covariates and treatment response clusters. Covariates selected for univariable modeling included age at diagnosis, female gender, body mass index (BMI), time to starting any dopaminergic therapy, drug class (i.e. L-dopa monotherapy, dopamine agonist monotherapy and L-dopa plus dopamine agonists), challenge test and total daily dose, education-adjusted MoCA score and MDS UPDRS part I – IV scores, with part III recorded in the on state. The following covariates with a P-value < 0.1 in at least in one group in univariable modeling were incorporated into the multivariable multinomial logistic regression model: age at diagnosis, female gender, BMI, time to starting dopaminergic therapy, drug class, challenge test and total daily dose, education-adjusted MoCA and MDS UPDRS II scores. MDS UPDRS III and IV were excluded from multivariable modeling as they closely correlate with percentage response. The multivariable model mutually adjusted for all included covariates. Results are reported in terms of multinomial odds ratio (95% CI, p-values). In a sensitivity analysis, we adjusted GMM for dosage to assess its impact on cluster identification. An additional analysis excluded observations beyond 9 years to evaluate the influence of late-stage dropout on clustering. All analyses were undertaken using R 4.3.0 software.

## Results

Of 423 cases enrolled, 6 (1.4%) had a change in diagnosis and were excluded, leaving 417 cases, of whom 336 (80.6%) had at least one challenge test. 287 of these (85.4%) underwent more than one challenge test. Of the 336 cases, 223 were male (66.4%) and 113 female (33.6%). At enrolment, the mean age was 61.2 years (SD 9.6) and mean disease duration 0.5 years (SD 0.5). Tables 1 and 2 present the demographic and clinical features of the cohort. A total of 1,527 challenge tests were performed, of which 2 were excluded as outliers, leaving 1,525 tests for analysis.

**TABLE 1:** Demographic and clinical characteristics in 336 patients with Parkinson’s disease, according to data-derived treatment response group.

| Variable | Response |  |  |  |
| --- | --- | --- | --- | --- |
|  | All | Modest | Excellent | Striking |
|  | N=336 | N=197 (58.6%) | N= 110 (32.7%) | N= 29 (8.7%) |
| **Age at diagnosis<sup>a</sup>** | 60.7 (9.6) | 61.9 (9.8) | 59.4 (8.8) | 57.4 (9.6) |
| **Disease duration at study entry** | 0.5 (0.5) | 0.6 (0.6) | 0.5 (0.5) | 0.4 (0.3) |
| **Male (%)** | 223 (66.4%) | 141 (71.6%) | 67 (60.9%) | 15 (51.7%) |
| **Baseline scoring** |  |  |  |  |
| **MDS UPDRS I scores** |  |  |  |  |
| - Total | 1.3 (1.6) | 1.2 (1.3) | 1.3 (1.8) | 1.6 (2.1) |
| - Anxiety >0 | 128 (38.1%) | 72 (36.5%) | 43 (39.1%) | 13 (44.8%) |
| - Depression >0 | 80 (23.8%) | 42 (21.3%) | 29 (26.4%) | 9 (31%) |
| **MDS UPDRS II total** | 5.7 (4.1) | 5.8 (4.1) | 5.4 (3.9) | 5.8 (4.5) |
| **MDS UPDRS III scores** |  |  |  |  |
| - Total | 20.8 (9.0) | 21.8 (9.2) | 19.8 (8.6) | 18.6 (8.4) |
| - Rest tremor score >0 | 239 (71.1%) | 133 (67.5%) | 86 (78.2%) | 20 (68.9%) |
| **MoCA total** | 27.1 (2.2) | 26.7 (2.3) | 27.3 (2.1) | 28.4 (1.3) |
| **BMI** | 27.1 (4.6) | 27.3 (4.8) | 27.1 (4.2) | 26.3 (4.4) |
| **Constipation** | 55 (16.4%) | 29 (14.7%) | 20 (18.2%) | 6 (20.7%) |
| **Education > 12 years** | 282 (80%) | 168 (85.3%) | 91 (82.7%) | 23 (79.3%) |
| **Duration of follow-up (years)** | 5.9 (2.0) | 6.0 (2.2) | 6.1 (1.8) | 5.2 (1.7) |
| **Progression scoring (annual rate of change)** |  |  |  |  |
| - MDS UPDRS II | 0.8 (0.7) | 0.9 (0.8) | 0.7 (0.7) | 0.6 (0.7) |
| - MDS UPDRS III | 1.8 (1.6) | 2.1 (1.6) | 1.4 (1.5) | 1.2 (1.7) |
| - MoCA | -0.06 (0.3) | -0.1 (0.3) | -0.02 (0.3) | 0.08 (0.2) |

BMI: body mass index; MDS UPDRS: Movement Disorder Society Unified
Parkinson's Disease Rating Scale; MoCA: Montreal Cognitive Assessment.
<sup>a</sup>Values are mean (standard deviation) unless otherwise indicated.

**TABLE 2:**
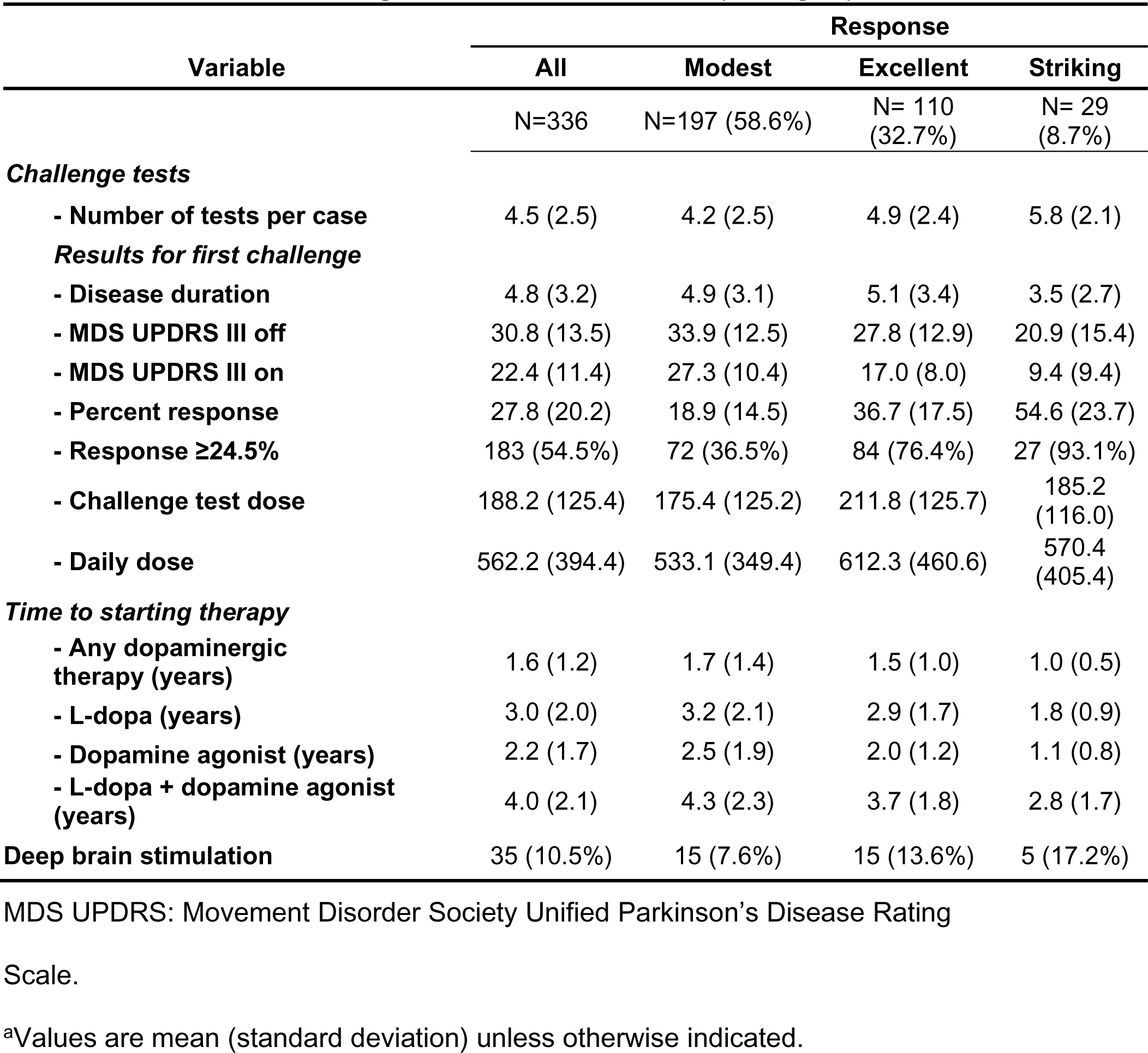
Challenge test results and dopaminergic therapy in 336 patients with Parkinson’s disease, according to data-derived treatment response group.

The 2 excluded challenge tests were from 2 separate cases; one test had a mismatch in the time of the challenge dose and the on and off scores, and the other had clearly inconsistent observations compared with their other scorings. The remaining challenge tests from these 2 cases were included in the analysis.

At 1-2 years after diagnosis, 48.0% of tests reached the 24.5% threshold for a definite response, but this proportion was higher for all subsequent 2-year time periods (between 51.1 and 74.3%) (Figure 1). In the 287 cases with more than one challenge test, an initially limited response became definite in 116 cases (40.4%) during a mean follow-up of 28.8 months (SD 19.7), whereas an initially definite response became limited in 72 cases (25.1%) over a mean follow-up of 22.9 months (SD 17.4). The remaining cases did not change category. In the 49 cases with only one challenge test, 25 (7.4% of 336 cases) had a limited response and 24 (7.1%) had a definite response. A limited response below the 24.5% threshold was more likely when motor scores were low, but was seen for all disease durations (Supplementary Figures 1 and 2). Duration of follow-up was similar between cases with an initially limited (55.4 months, SD 35.7) and initially definite response (50.0 months, SD 34.9). 85.6% of initially limited responders and 85.2% of initially definite responders went on to have 1 or more challenge tests.

**FIG. 1.**
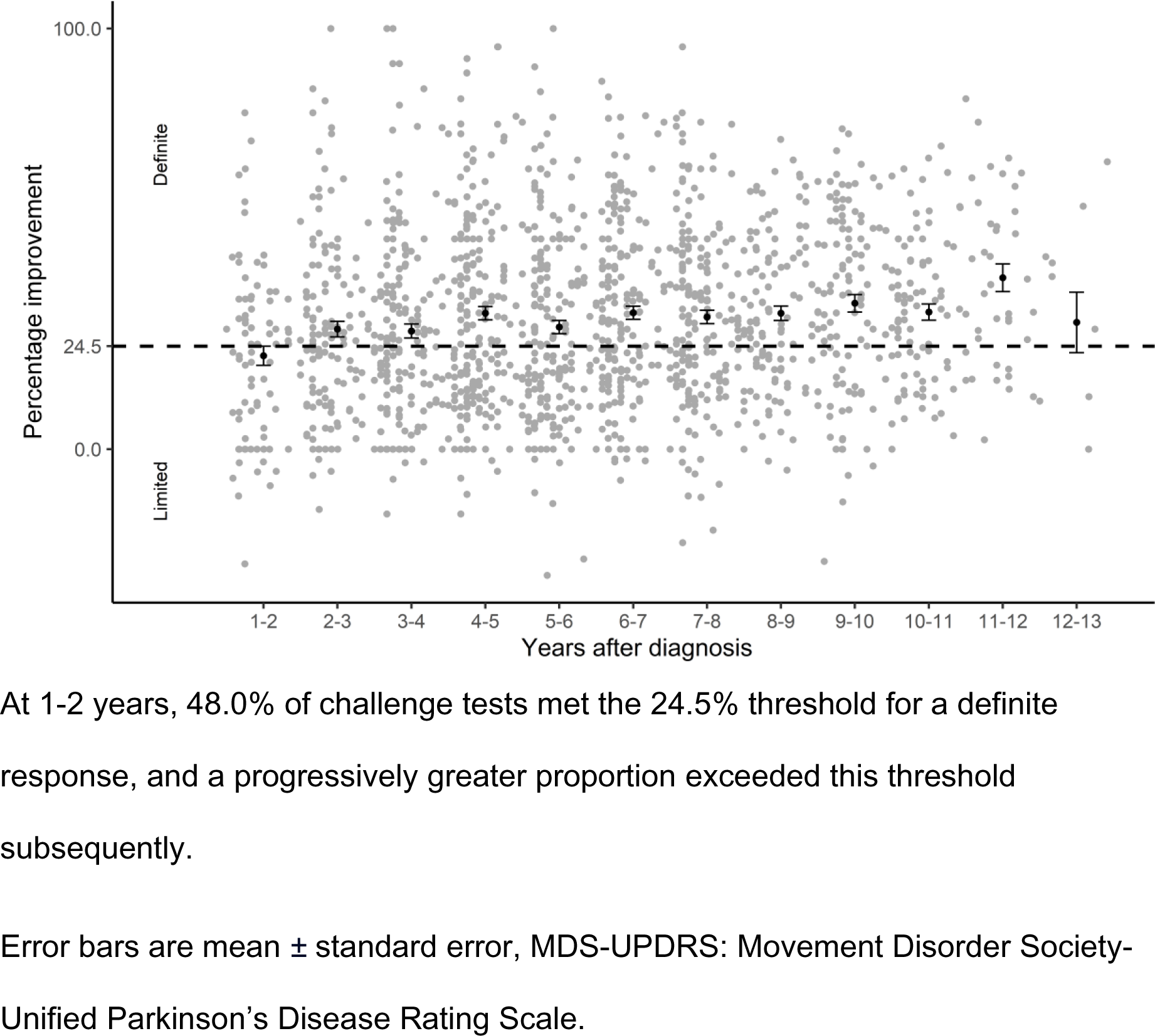
Percentage change in MDS-UPDRS part III scores in 1525 challenge tests in 336 cases of Parkinson’s disease.

Both the challenge test and total daily dose of dopaminergic medication increased over time (Table 3). Dopaminergic drug class affected percentage responsiveness: a significantly greater percentage response was seen with L-dopa compared to a dopamine agonist alone (P < 0.0001). A similar proportion had a definite response when a combination of drug classes was used compared to L-dopa alone (P = 0.37).

**TABLE 3:**
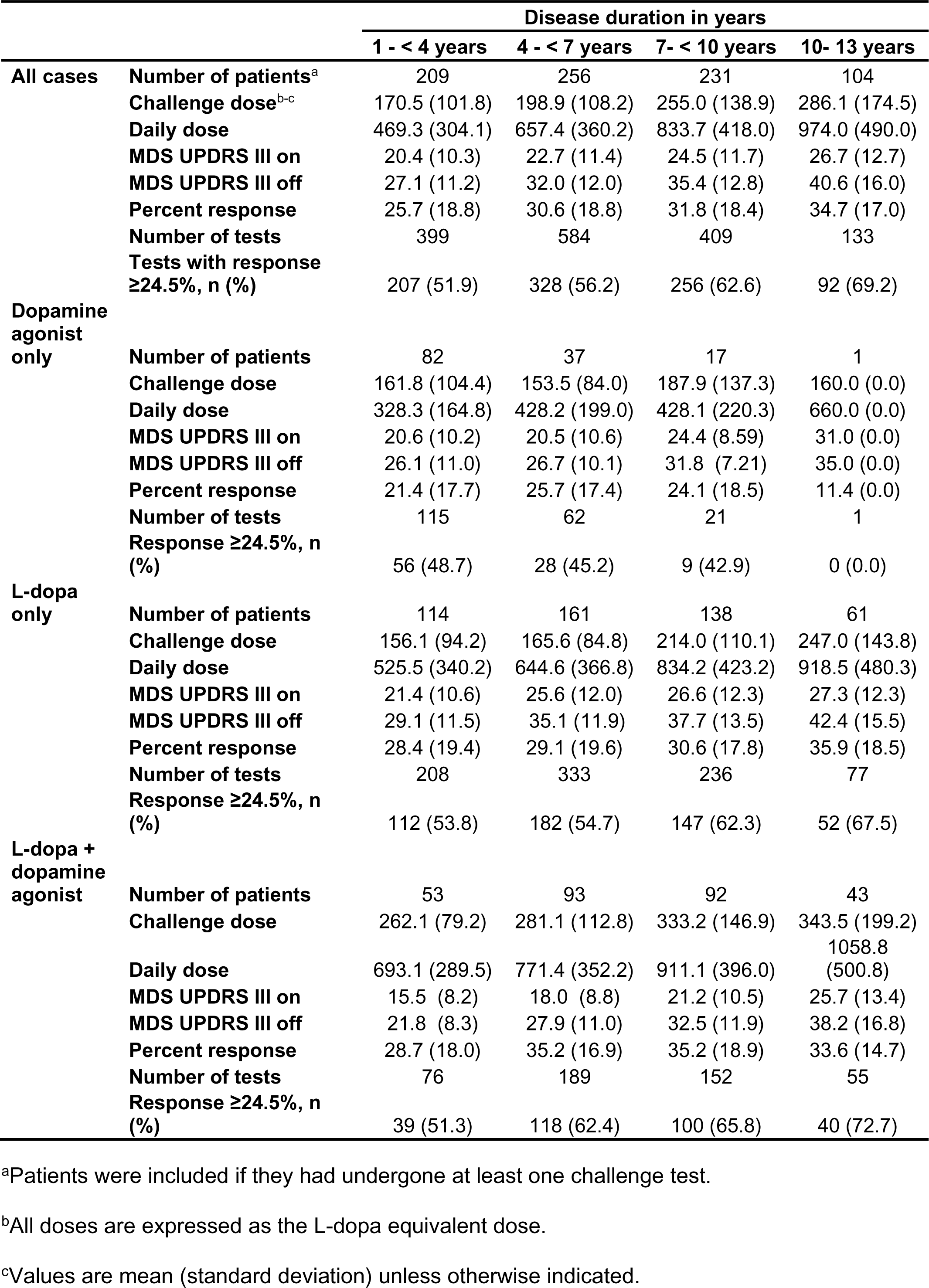
Challenge test responses according to drug class and disease duration.

|  |  | Disease duration in years |  |  |  |
| --- | --- | --- | --- | --- | --- |
|  |  | 1 - < 4 years | 4 - < 7 years | 7- < 10 years | 10- 13 years |
| **All cases** | **Number of patients<sup>a</sup>** | 209 | 256 | 231 | 104 |
|  | **Challenge dose<sup>b-c</sup>** | 170.5 (101.8) | 198.9 (108.2) | 255.0 (138.9) | 286.1 (174.5) |
|  | **Daily dose** | 469.3 (304.1) | 657.4 (360.2) | 833.7 (418.0) | 974.0 (490.0) |
|  | **MDS UPDRS III on** | 20.4 (10.3) | 22.7 (11.4) | 24.5 (11.7) | 26.7 (12.7) |
|  | **MDS UPDRS III off** | 27.1 (11.2) | 32.0 (12.0) | 35.4 (12.8) | 40.6 (16.0) |
|  | **Percent response** | 25.7 (18.8) | 30.6 (18.8) | 31.8 (18.4) | 34.7 (17.0) |
|  | **Number of tests** | 399 | 584 | 409 | 133 |
|  | **Tests with response $\geq 24.5\%$, n (%)** | 207 (51.9) | 328 (56.2) | 256 (62.6) | 92 (69.2) |
| **Dopamine agonist only** | **Number of patients** | 82 | 37 | 17 | 1 |
|  | **Challenge dose** | 161.8 (104.4) | 153.5 (84.0) | 187.9 (137.3) | 160.0 (0.0) |
|  | **Daily dose** | 328.3 (164.8) | 428.2 (199.0) | 428.1 (220.3) | 660.0 (0.0) |
|  | **MDS UPDRS III on** | 20.6 (10.2) | 20.5 (10.6) | 24.4 (8.59) | 31.0 (0.0) |
|  | **MDS UPDRS III off** | 26.1 (11.0) | 26.7 (10.1) | 31.8 (7.21) | 35.0 (0.0) |
|  | **Percent response** | 21.4 (17.7) | 25.7 (17.4) | 24.1 (18.5) | 11.4 (0.0) |
|  | **Number of tests** | 115 | 62 | 21 | 1 |
|  | **Response $\geq 24.5\%$, n (%)** | 56 (48.7) | 28 (45.2) | 9 (42.9) | 0 (0.0) |
| **L-dopa only** | **Number of patients** | 114 | 161 | 138 | 61 |
|  | **Challenge dose** | 156.1 (94.2) | 165.6 (84.8) | 214.0 (110.1) | 247.0 (143.8) |
|  | **Daily dose** | 525.5 (340.2) | 644.6 (366.8) | 834.2 (423.2) | 918.5 (480.3) |
|  | **MDS UPDRS III on** | 21.4 (10.6) | 25.6 (12.0) | 26.6 (12.3) | 27.3 (12.3) |
|  | **MDS UPDRS III off** | 29.1 (11.5) | 35.1 (11.9) | 37.7 (13.5) | 42.4 (15.5) |
|  | **Percent response** | 28.4 (19.4) | 29.1 (19.6) | 30.6 (17.8) | 35.9 (18.5) |
|  | **Number of tests** | 208 | 333 | 236 | 77 |
|  | **Response $\geq 24.5\%$, n (%)** | 112 (53.8) | 182 (54.7) | 147 (62.3) | 52 (67.5) |
| **L-dopa + dopamine agonist** | **Number of patients** | 53 | 93 | 92 | 43 |
|  | **Challenge dose** | 262.1 (79.2) | 281.1 (112.8) | 333.2 (146.9) | 343.5 (199.2) |
|  | **Daily dose** | 693.1 (289.5) | 771.4 (352.2) | 911.1 (396.0) | 1058.8 (500.8) |
|  | **MDS UPDRS III on** | 15.5 (8.2) | 18.0 (8.8) | 21.2 (10.5) | 25.7 (13.4) |
|  | **MDS UPDRS III off** | 21.8 (8.3) | 27.9 (11.0) | 32.5 (11.9) | 38.2 (16.8) |
|  | **Percent response** | 28.7 (18.0) | 35.2 (16.9) | 35.2 (18.9) | 33.6 (14.7) |
|  | **Number of tests** | 76 | 189 | 152 | 55 |
|  | **Response $\geq 24.5\%$, n (%)** | 39 (51.3) | 118 (62.4) | 100 (65.8) | 40 (72.7) |

<sup>a</sup>Patients were included if they had undergone at least one challenge test.
<sup>b</sup>All doses are expressed as the L-dopa equivalent dose.
<sup>c</sup>Values are mean (standard deviation) unless otherwise indicated.

### Cluster identification from modeling

In GMM, we observed identical SABIC values for 2 or 3 clusters (13235) and similar entropy (0.67 and 0.61, respectively) and AIC values (13226.8 and 13223.5, respectively). After truncating the data at 9 years and repeating GMM, SABIC and AIC were lower for three (SABIC=11210.6 and AIC=11199.9) compared to two clusters (SABIC=11212.1, AIC=11203.7) with similar entropy values (3 clusters= 0.63, 2 clusters=0.68). As lower SABIC and AIC values are preferred, a 3-cluster model was favored (Figure 2, Supplementary Figure 3). Entropy provides insight into the "overlap" between clusters, reflecting uncertainty about which cluster some individuals belong to. Despite slightly higher entropy values for two clusters (0.67 for 2 clusters, 0.61 for 3 clusters), the posterior probability of belonging to the assigned class in the 3-cluster model remains high at a 50% threshold (Supplementary Figure 4, Supplementary Table 1). The 3 cluster model was additionally considered more clinically informative and a better reflection of clinical experience (Figure 2, Supplementary Figure 3).

**FIG. 2.**
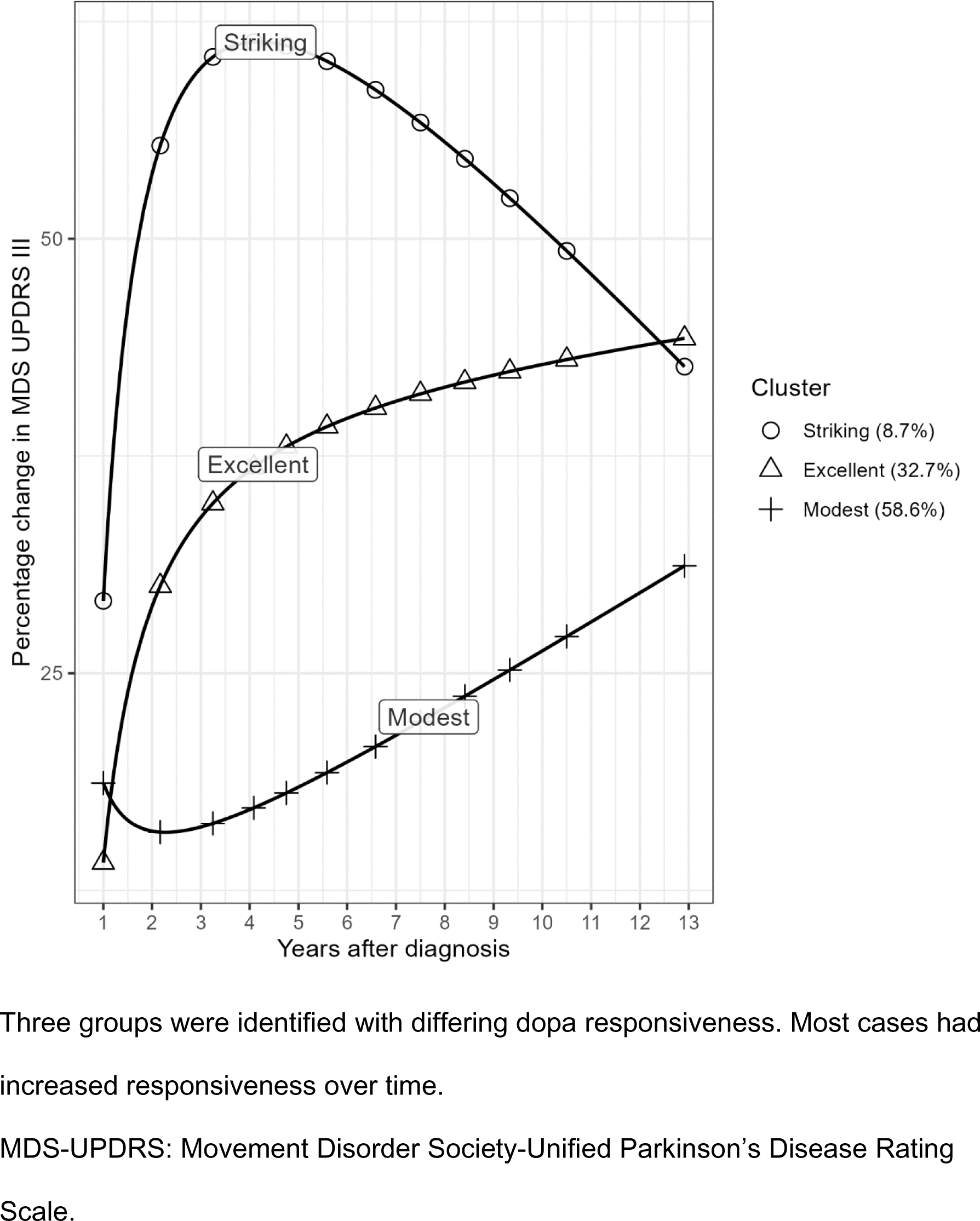
Trajectory of dopaminergic responsiveness over time according to data- driven cluster categorization.

The following labels were used to describe these 3 clusters: (i) Striking response in 29 cases (8.7%), (ii) Excellent response in 110 cases (32.7%), and (iii) Modest response in 197 cases (58.6%). Sensitivity analysis excluding data beyond 9 years identified the same 3 clusters (Supplementary Figure 5), indicating that attrition did not influence cluster identification, allocation, or trajectory. Loss to follow up rates were similar in striking (20.7%) and modest (23.4%) responders, and most patients were followed for the study duration (76.6% to 84.5% across the 3 clusters) (Supplementary Table 2).

GMM trajectories were additionally adjusted for challenge test dose and to visualize this modelling approach we used a reference dose of 200mg (Supplementary Figure 6). Dose varied by group and time (Supplementary Table 3), most notably in striking and excellent responders. When comparing GMM with and without dose adjustment, modest and excellent responders had very similar trajectories. For the striking group there was a greater reduction in response in later disease, suggesting that higher doses contributed to the better response in these cases.

In univariable modeling, striking and excellent responders were significantly more likely to be female, younger, have lower MDS UPDRS II and III ‘on’ scores, higher MDS UPDRS IV and MoCA scores, and have a higher challenge dose compared with modest responders (Figure 3). A striking response was associated with significantly lower BMI. In the multivariable model (Figure 4), striking responders started dopaminergic therapy significantly earlier (P = 0.02), were less likely to be on dopamine agonist monotherapy (P = 0.01) and had significantly better cognition (P < 0.01) and MDS UPDRS II scores (P = 0.01). Excellent responders were significantly more likely to be on a combination of drug classes (P < 0.01) and had a higher challenge dose (P = 0.03).

**FIG. 3.**
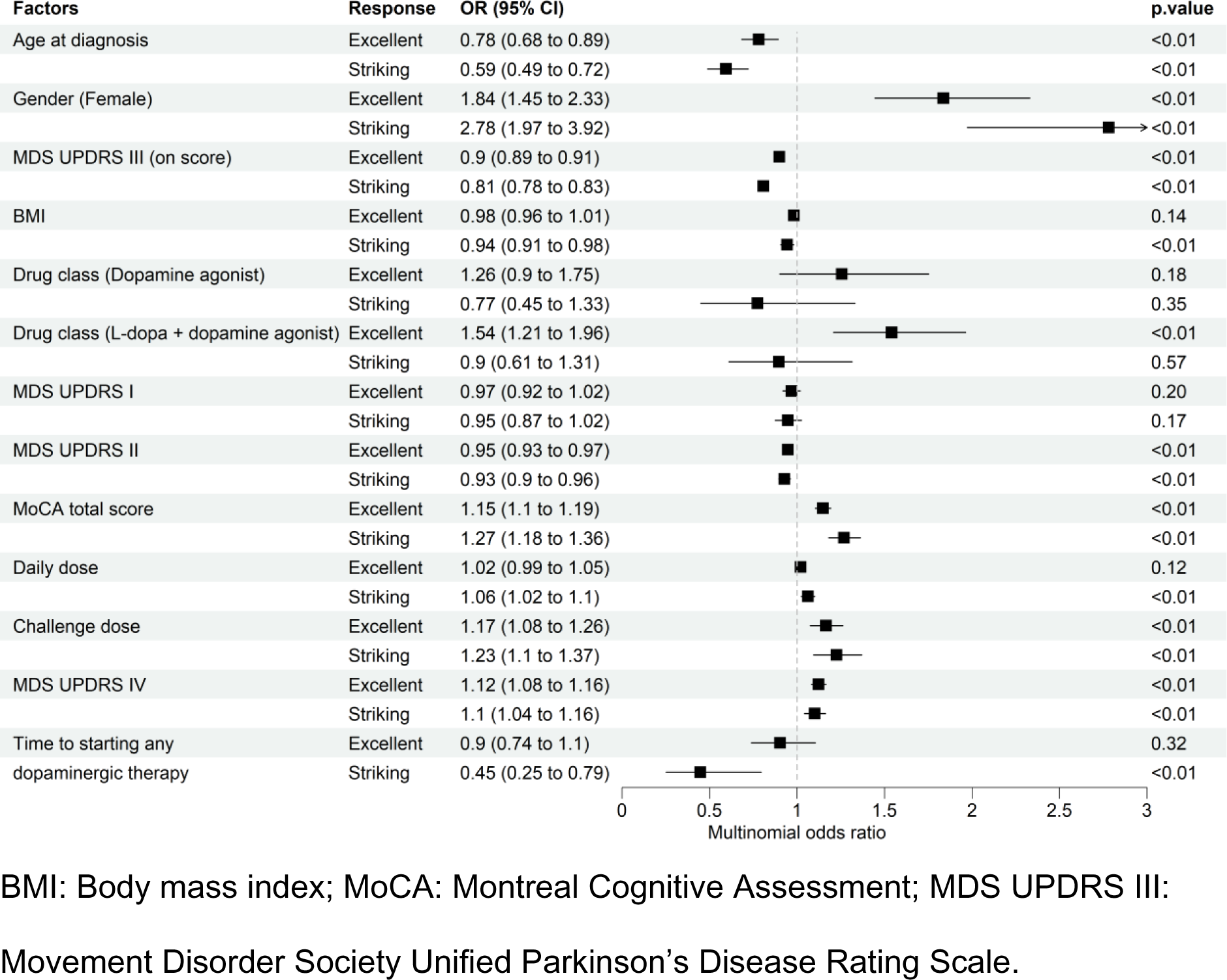
Odds ratios from univariable modeling of factors associated with dopa responsiveness.

**FIG. 4.**
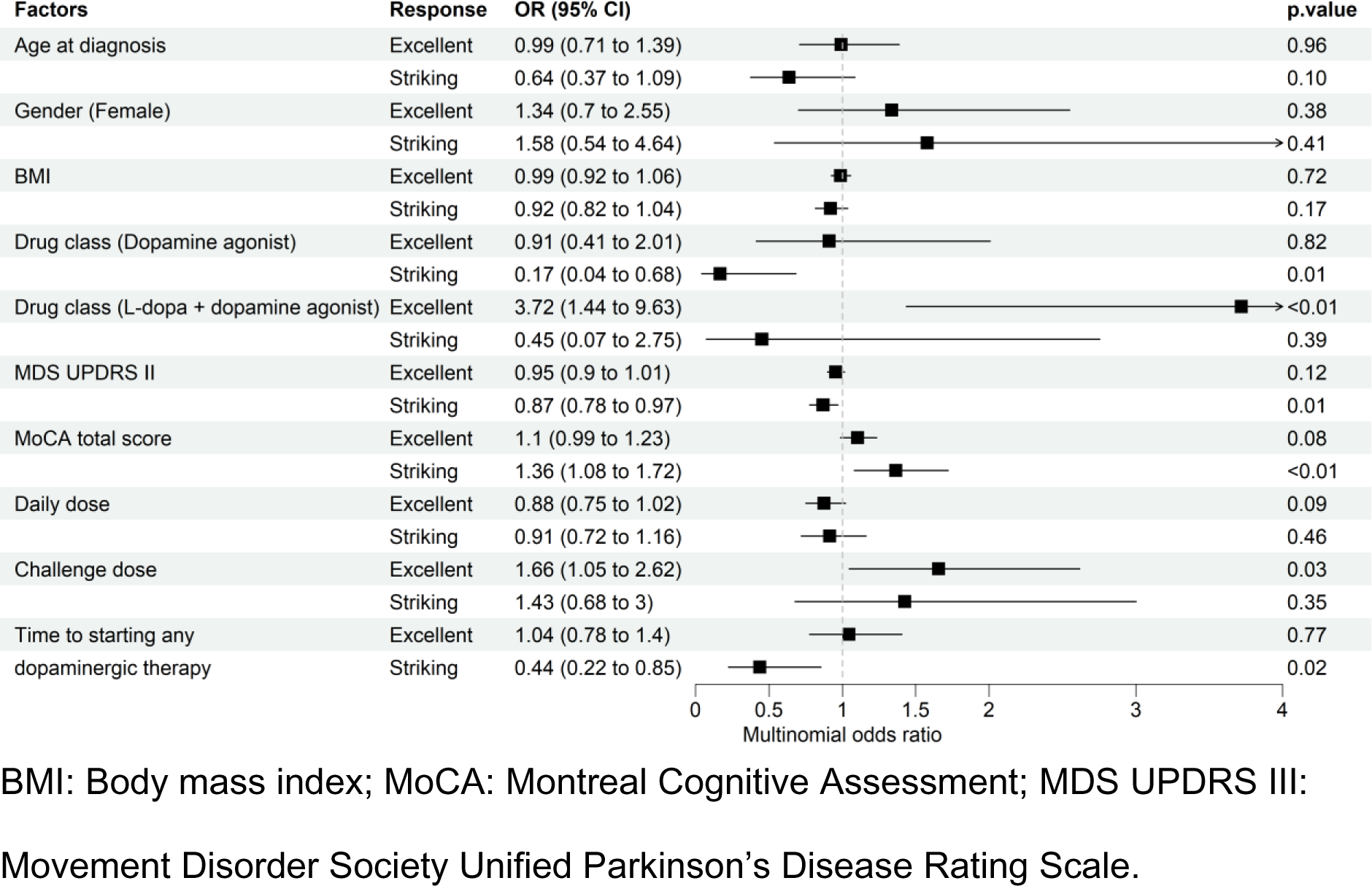
Odds ratios from multivariable modeling of factors associated with dopa responsiveness.

We then examined how challenge test responses and motor complications differed according to cluster and disease duration (Supplementary Table 4). Most differences between clusters persisted over time, though in the case of daily dose and MDS UPDRS IV the differences between clusters increased over time and then weakened (or were consistent with chance) at 10-13 years of disease, which had the least power. Striking and excellent responders were significantly more likely to have a definite treatment response than modest responders, and had lower motor ‘on’ and ‘off’ scores (Figure 5) and higher MDS UPDRS IV scores (Table 2).

**FIG. 5.**
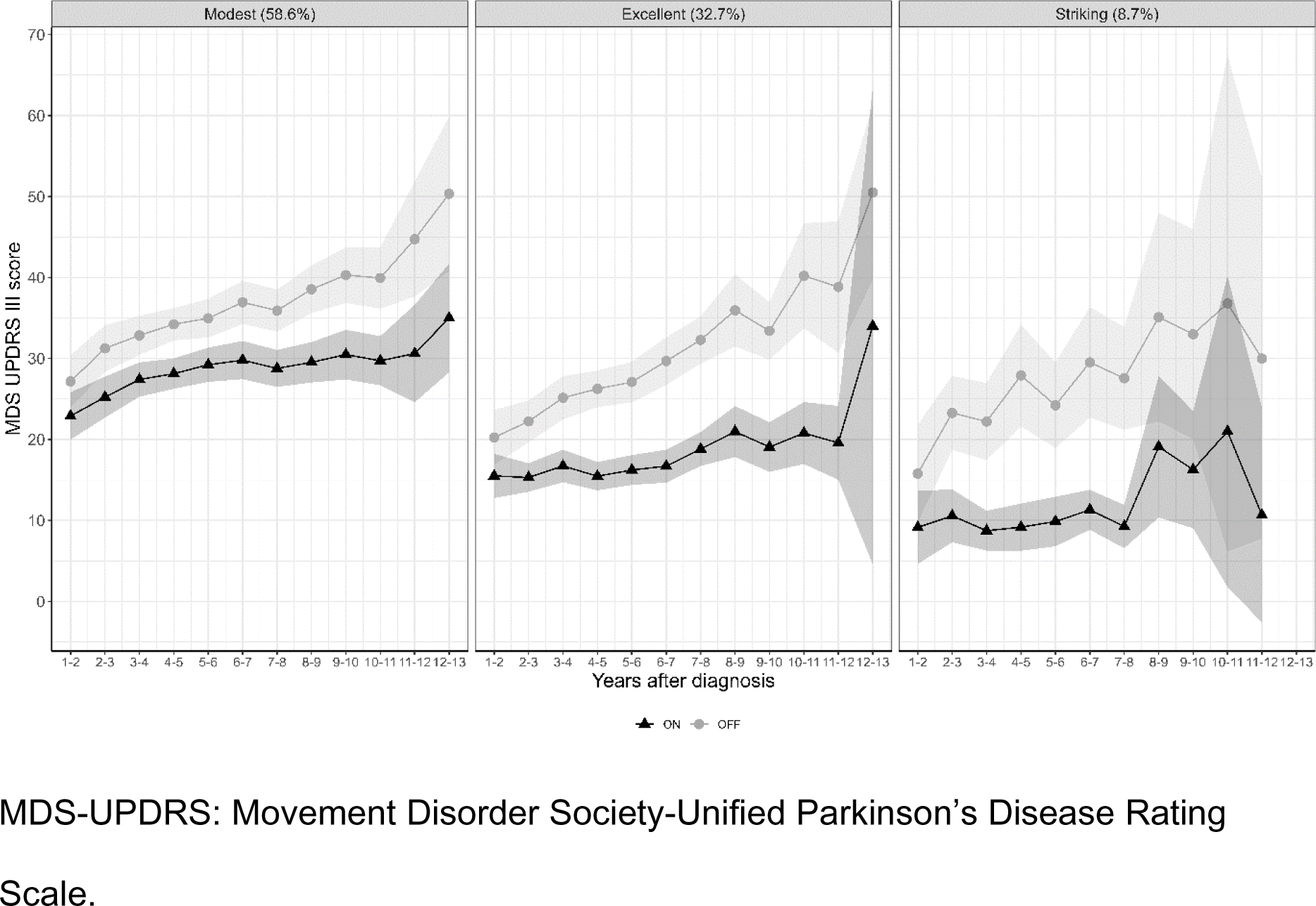
Motor scores in the off and on state in 336 cases of Parkinson’s disease, categorized by test result.

### Subjective grading - individual patients

Subjective assessments of the L-dopa response were available in 372 of 417 cases (89.2%), of which 359 (96.5%) had a positive L-dopa response and 13 (3.5%) had ‘little or no response’. In the 359 with a positive response, grading was available for 209 cases and was marked in 196 (93.8%), and moderate in 13 (6.2%).

### Subjective grading - individual challenge tests

Clinicians subjectively graded the L-dopa response in 1,366 of 1,525 challenge tests (89.6%); a response was graded as subjectively present in 1,335 (97.7%) and ‘little or no response’ was recorded on 31 occasions (2.3%). The measured response on the challenge test was 32.2% (SD 21.2) when subjectively considered present versus 24.4% (SD 21.5) when tests were subjectively considered to show ‘little or no response’. For cases when the L-dopa response was graded, based on the updated protocol, a marked response corresponded to a measured response of 36.1% (SD 19.0) for 225 tests, while a moderate response corresponded to a measured response of 28.9% (SD 21.2) for 9 tests.

## Discussion

The current report is, to our knowledge, the most detailed analysis of the treatment response in PD, covering cases from diagnosis to 13 years later, and assessing the response to both dopamine agonists and L-dopa. Our novel finding is that there are three distinct patterns of dopaminergic responsiveness in PD. A striking response, exceeding 50% improvement, was seen in just under 10% of cases. This degree of improvement was previously reported in a subset of PD cases selected for deep brain stimulation surgery^8^. An excellent response was seen in around one third, and a modest response in just over half of cases. More modest L-dopa responses have also been reported previously during the first year of treatment in the *ELLDOPA* study, including in patients with a daily dose of 600mg L-dopa^2^.

Our finding was obtained from a primarily data-driven analysis of measured motor scores, without the influence of subjective assessments of the treatment response. Previous observational cohort studies found that on and off motor scores increased^9–11, 13, 24^, but the response amplitude was either conserved^13, 24^ or increased over time^9–11^. Several of these studies were performed in specific PD subgroups with motor fluctuations^13, 24^, involved the superseded Modified Webster scale^9–11, 13^, and cohorts that were too small to undertake a detailed data-driven cluster analysis^9–13, 24^.

Our cluster analysis confirms that there is a progressive increase in the difference between ‘on’ and ‘off’ scores over time, both for patients with an excellent and modest response, which represent over 90% of PD cases. In early disease, the lower doses used make it difficult to determine if there is a definite treatment response. However, over time, progressively larger proportions of PD cases fulfil the defined threshold for definite dopaminergic responsiveness. This has significant clinical implications, as many PD cases who initially do not fulfil supportive diagnostic criteria for a positive response to dopaminergic therapy^1^ will do so later in their disease course. In the 10% of cases with an initial striking response, a further increment was less obvious, perhaps because the ceiling of response had been reached. These differences are not simply explained by higher doses being associated with greater improvement (Supplementary Table 4). From challenge test doses of around 180-220mg in early disease, striking cases had over 50% improvement, whereas excellent responders had 30 to 40% improvement. Modest responders, conversely, had less than 30% improvement from the higher doses (230-260mg) reached in later disease, thereby challenging the concept that greater improvement would be observed at higher doses.

Compared with prior reports, a greater proportion of cases were subjectively graded as having a marked response (93.8%), but a similar proportion were identified as poor responders (3.5%). In one study of 69 cases, the response was excellent in 29%, good in 54%, moderate in 13%, and nil-to-poor in 4%^4^. In another study of 182 cases, 69% had a marked response, 22% were graded as moderate, and 9% had no response^5^. Both of these reports extracted data from case record review, which may explain the differences from the *PPMI* study, where grading was assessed prospectively. On-off motor scoring using challenge tests in another cohort found that 61% of 1007 cases had a definite response while 39% had a limited response, at 3.4 years^3^ of disease, which is somewhat higher than the current findings where 51% had a definite response and 49% had a limited response 3-4 years after diagnosis. The lower response in the current analysis may be explained in part by including challenge tests with dopamine agonists, rather than only L-dopa^3^.

There were mismatches between the subjective interpretation of the L-dopa response and the measured scores, for both the absence of an L-dopa response where there was a definite measured response, and vice versa. Accordingly a PD diagnosis should not be excluded based on a subjective assessment of ‘little or no response’ to L-dopa, particularly in early disease, which is in agreement with the conclusions from the *ELLDOPA* study^2^.

Our univariable model results are partially consistent with previous analyses of the demographic and clinical factors associated with L-dopa responsiveness. Our cases with greater dopaminergic responsiveness were significantly younger and had lower motor scores, as reported in a study of 1007 cases undergoing challenge tests^2, 3^, but there was no age association in another study assessing longer-term treatment responses^2^. In the current analysis, female patients had a better response than male patients, which compares to no significant gender difference in one study^3^ and some evidence of a better response in male patients in another^2^. However, in the multivariable model, both age and gender were consistent with chance. Several drug factors (higher doses, drug class, and starting treatment earlier), and better cognition and activity of daily living scores remained predictors, which were not included in prior studies^2, 3^. While the methodology is clearly different for challenge tests versus outpatient derived treatment response, an overall interpretation is that age and gender are not key factors related to the treatment response in PD.

GMM was chosen as one of the most sophisticated modeling approaches to identify subgroups with different trajectories^25–27^. While we considered group-based trajectory modeling (GBTM), GMM provides a better fit for the data, allows for within-cluster variability and most closely approximates the true group trajectories underlying the data^28^.

The similar entropy values of the different models, however, does indicate a degree of uncertainty as to which cluster some cases may belong to, which is a limitation of this study. A further limitation is the lack of pathological confirmation of PD. While benign disorders were excluded by confirming presynaptic dopamine depletion on functional imaging, and an expert panel excluded cases with other degenerative parkinsonian disorders, it is possible that some included cases do not have PD.

Motor scores were lower at baseline in striking and excellent responders than in modest responders, likely reflecting greater dopaminergic responsiveness. As percentage response was calculated by off score – on score / off score, this could have had a potential floor effect in modest responders. Despite this, percentage change was still considered preferable to absolute change, as it is defined in the MDS diagnostic criteria^1^ and is consistent with the methodological approach of previous studies^2, 3^. A small subset of our cohort (n = 35, 10.5%) underwent DBS. Although striking and excellent responders were more likely to have DBS (striking 17.2% and excellent 13.6% vs. modest responders 7.6%), this is likely to be explained by case selection for DBS based on the degree of dopa responsiveness. It is unlikely that DBS confounded the assessment of dopaminergic responsiveness, as it was generally performed in late disease, was switched off for off scores, and a response to challenge tests would still be expected after DBS implantation. Finally, although some patients were lost to follow-up during the later stages of disease, rates of drop out were similar between striking and modest responders. Moreover, the sensitivity analysis identified the same three treatment response groups with no reduction in the proportion with modest responses.

In conclusion, we have identified 3 distinct types of dopaminergic medication response in Parkinson’s using a data-driven analysis. Drug factors appear to contribute more than demographic factors to these groupings. As the proportion of PD cases fulfilling criteria for dopa responsiveness progressively increases over time, the initial response to treatment may be unreliable, which is relevant to the diagnostic process.

## Data Availability

All data produced in the present work are contained in the manuscript.

## Acknowledgements

We would like to thank Dr Tanja Zerenner at the Population Health Sciences Institute at the University of Bristol for her assistance with coding issues.

This analysis uses data openly available from *PPMI* (Tier 1), obtained in November 2022 from the *Parkinson’s Progression Markers Initiative* (*PPMI*) database (www.ppmi-info.org/access-data-specimens/download-data), RRID:SCR_006431. For up-to-date information on the study, visit www.ppmi-info.org.

## Author Roles

1. Research project: A. Conception, B. Organization, C. Execution;

2. Statistical analysis: A. Design, B. Execution, C. Review and Critique;

3. Manuscript Preparation: A. Writing of the first draft, B. Review and Critique.

SEG: 1B, 1C, 2A, 2B, 2C, 3A, 3B.

AN: 1B, 1C, 2A, 2B, 2C, 3A, 3B MAL: 2A, 2C, 3B

VM: 2C, 3B YBS: 2A, 2C, 3B

KAG, DGG: 1A, 1B, 1C, 2A, 2C, 3A, 3B

## Disclosures

### Financial disclosures for the previous 12 months

MAL received consultancy fees for advising on a secondary analysis of an RCT sponsored by North Bristol NHS trust.

VM has received funding for travel to a conference for BIAL and lecture fees from BIAL.

KAG has received consultancy fees from Parkinson’s UK.

YBS has received consultancy fees from Human Centric DD Ltd and Parkinson’s UK.

DGG has received consultancy fees from Parkinson’s UK and NeuroClin Glasgow.

SEG: none; AN: none.

### Ethical Compliance Statement

*PPMI* was conducted following approval of the local ethics committees of the participating sites, in accordance with national legislation and the Declaration of Helsinki. Written informed consent was obtained from each participant prior to enrollment. We confirm that we have read the Journal’s position on issues involved in ethical publication and affirm that this work is consistent with those guidelines.

---

**Supplementary Figure 1:**
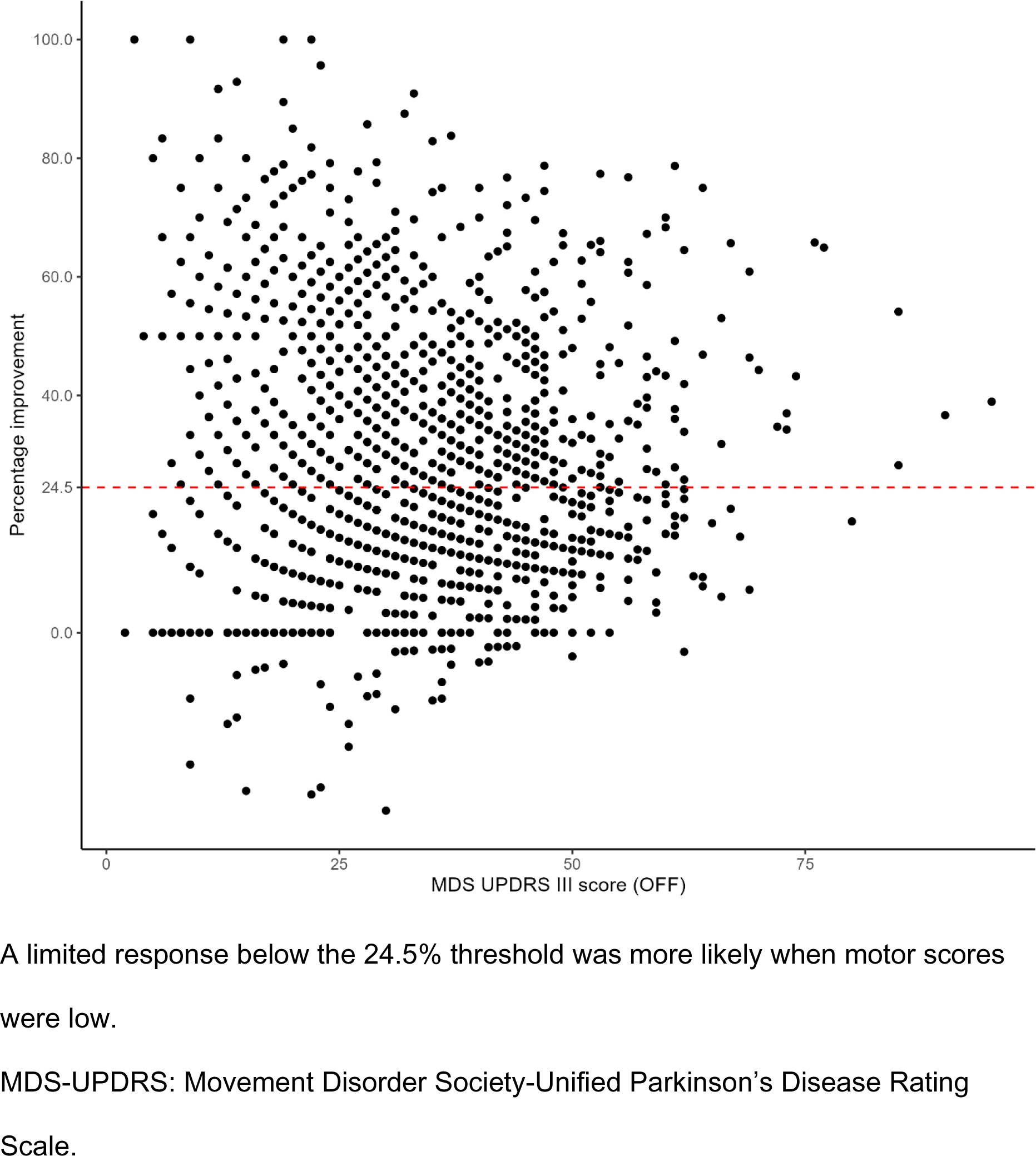
Scatterplot of percentage improvement versus ‘off’ motor scores.

**Supplementary Figure 2:**
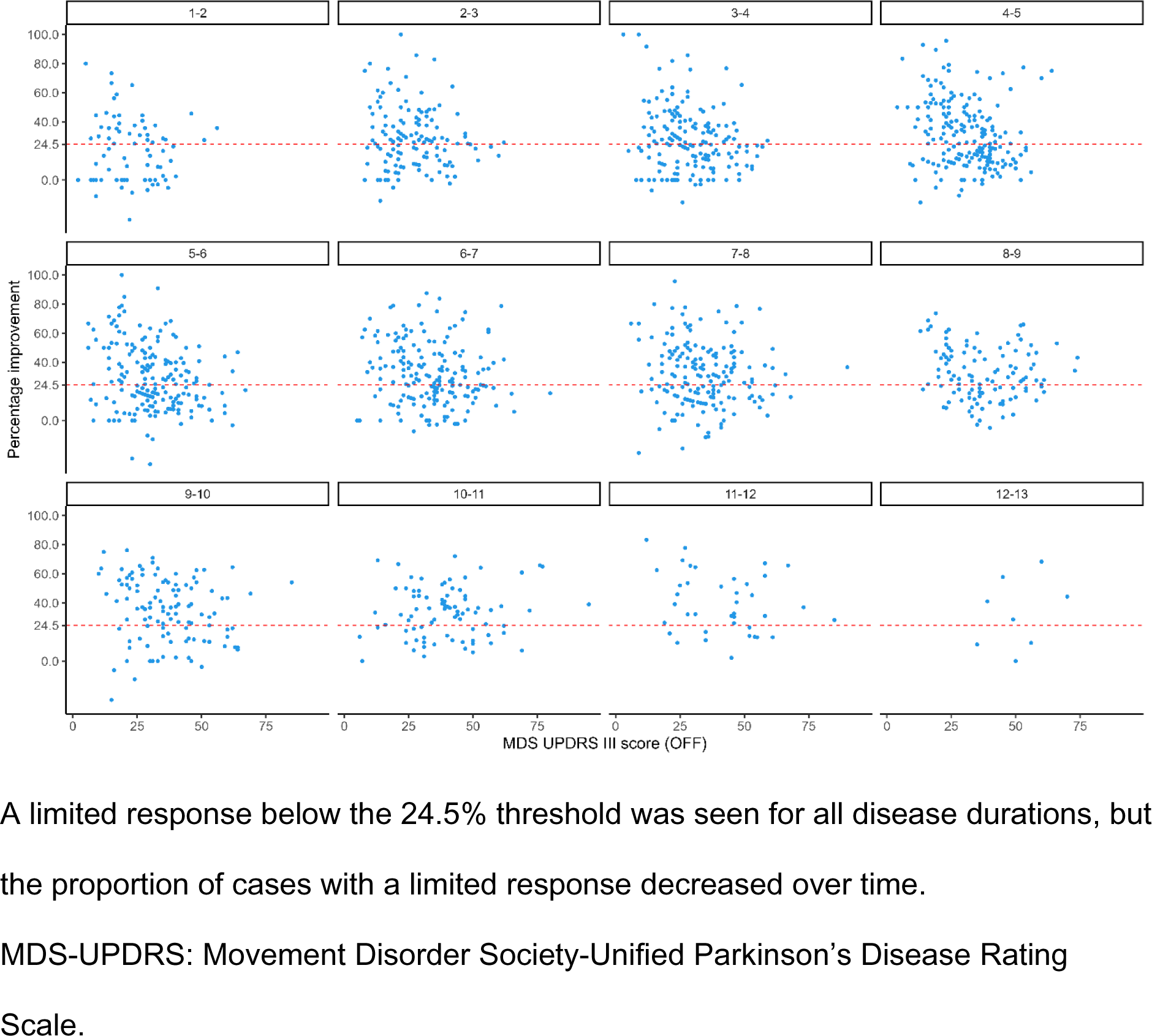
Scatterplot of percentage improvement versus ‘off’ motor scores, according to disease duration.

**Supplementary Figure 3:**
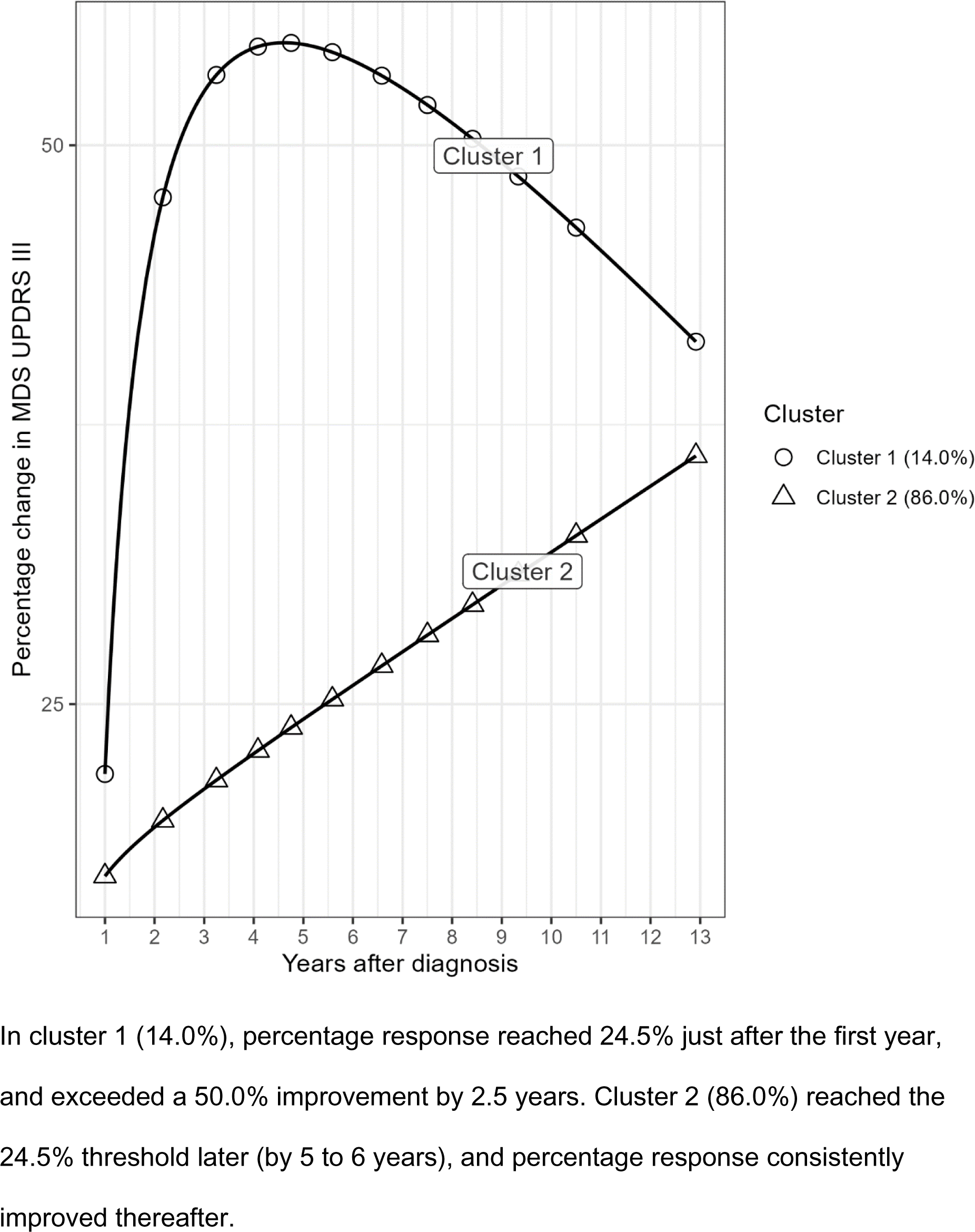
GMM using 2 clusters.

**Supplementary Figure 4:**
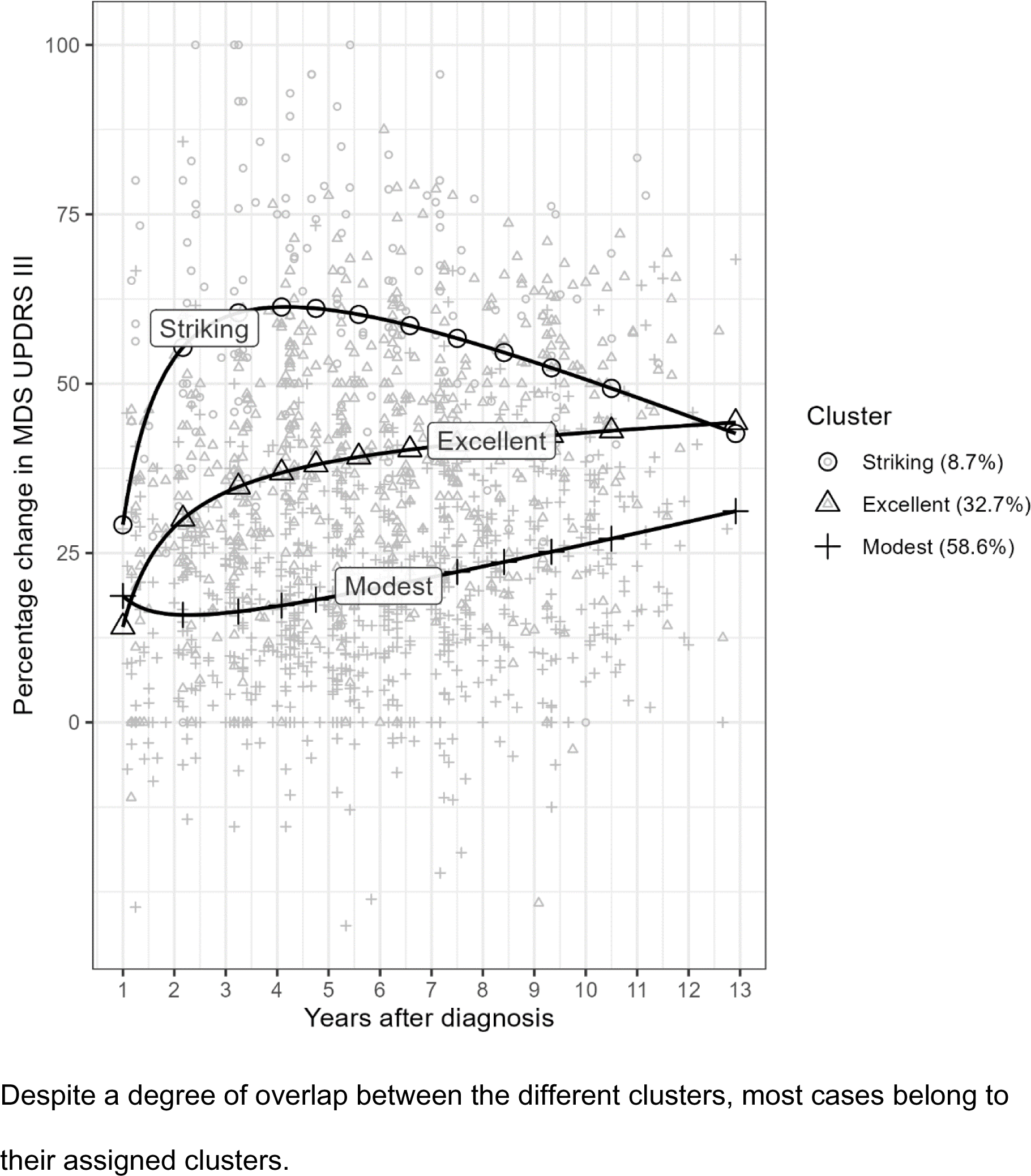
GMM with raw data points, shape-coded according to cluster.

**Supplementary Figure 5:**
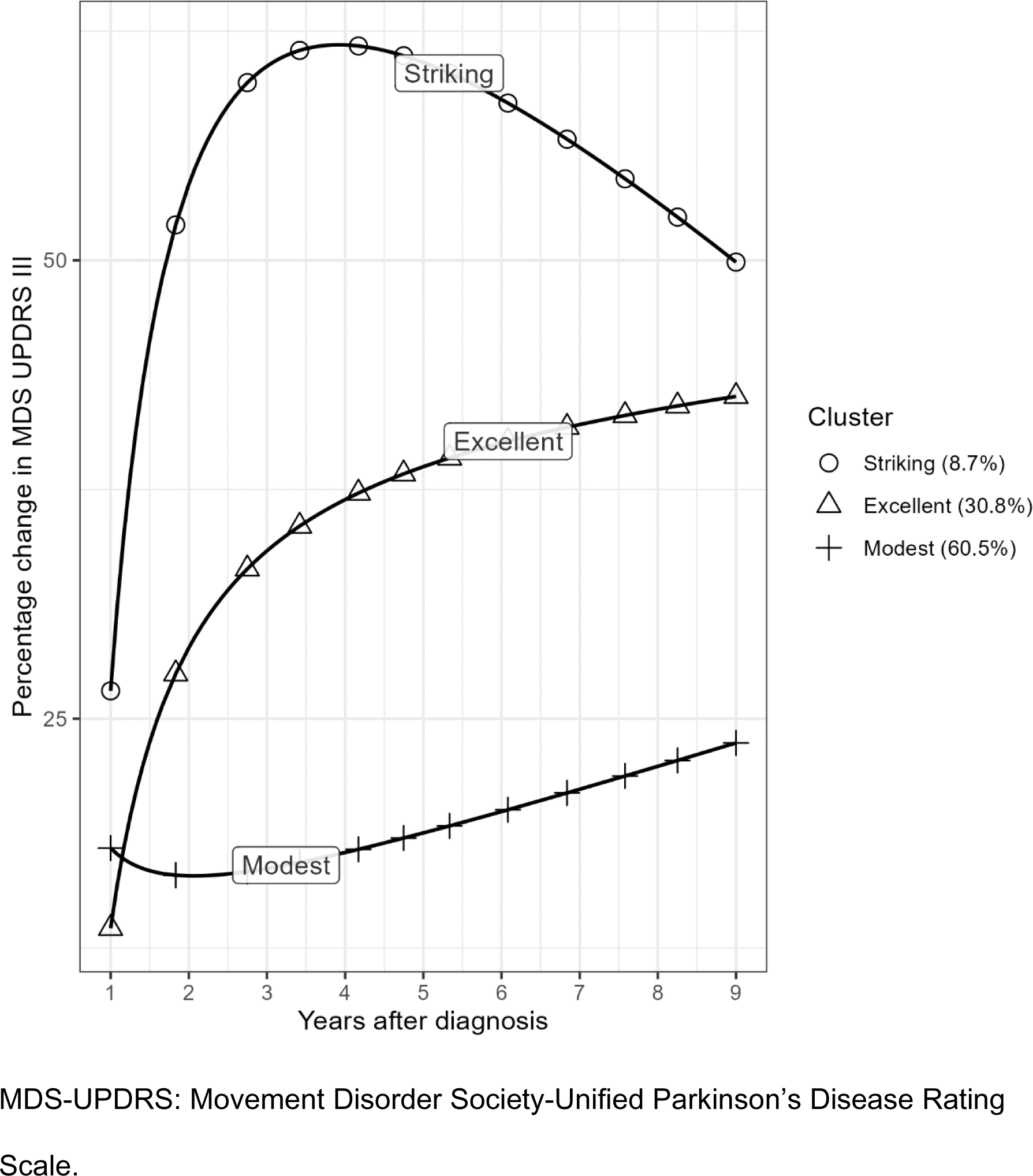
Sensitivity analysis excluding data beyond 9 years. GMM identified the same three treatment response groups, with no indication of selective dropout of cases from the modest treatment response group.

**Supplementary Figure 6:**
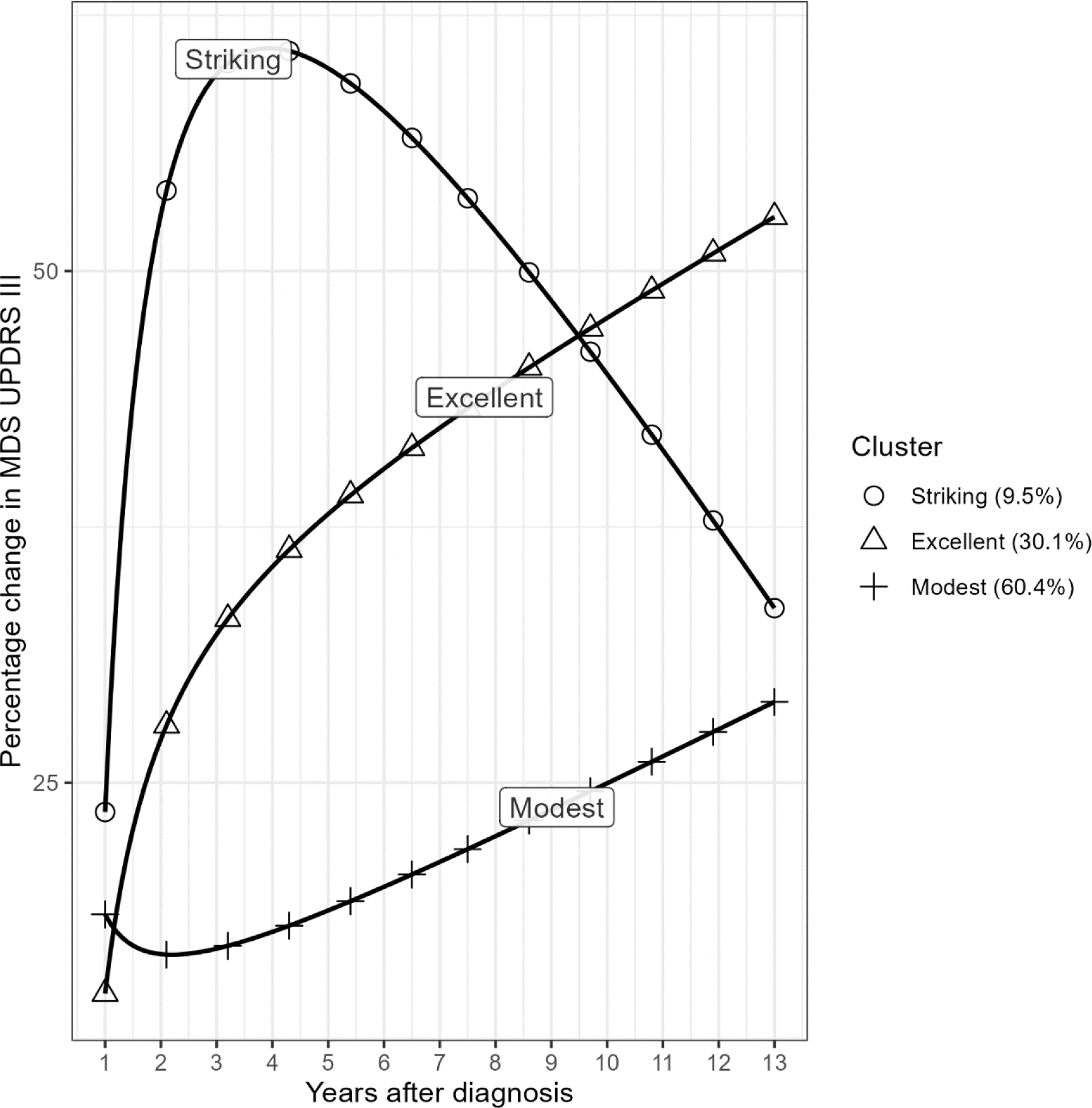
GMM adjusted for dose using a 200mg reference dose. When comparing GMM with and without dose adjustment, modest and excellent responders have very similar trajectories. Whilst the trajectories for striking responders differ in later disease, this partly reflects the fixed 200mg dose used in the adjusted model.

