## Supplementary Table 1 for "Dopa responsiveness in Parkinson’s disease"

***SUPPLEMENTARY TABLE 1:*** Probability of belonging to the assigned class based on entropy values

| **Probability** | **Class 1 (n = 197) Modest** | **Class 2 (n = 29) Striking** | **Class 3 (n = 110) Excellent** |
|---|---|---|---|
| 0.5 | 195 (98.9%) | 29 (100%) | 106 (96.3%) |
| 0.6 | 172 (87.3%) | 26 (89.6%) | 76 (69.1%) |
| 0.7 | 158 (80.2%) | 23 (79.3%) | 58 (52.7%) |
| 0.8 | 130 (65.9%) | 19 (65.5%) | 45 (40.9%) |
| 0.9 | 93 (47.2%) | 18 (62.1%) | 28 (25.4%) |
