## Supplementary Table 2 for "Dopa responsiveness in Parkinson’s disease"

***SUPPLEMENTARY TABLE 2:*** Analysis of follow-up across the three treatment-response groups

| **Treatment response-groups** | **Loss to follow-up** | |
|---|---|---|
| | **Yes** | **No** |
| **Striking (n = 29)** | 6 (20.7%) | 23 (79.3%) |
| **Excellent (n = 110)** | 17 (15.5%) | 93 (84.5%) |
| **Modest (n = 197)** | 46 (23.4%) | 151 (76.6%) |

Results are reported as frequency (%).
