## Supplementary Table 3 for "Dopa responsiveness in Parkinson’s disease"

***SUPPLEMENTARY TABLE 3:*** Dosage Follow-Up Analysis: A Comparison Across Three Treatment-Response Groups

| **Time bucket (years)** | **Treatment response-groups** | | |
|---|---|---|---|
| | **Striking (n = 29)** | **Excellent (n = 110)** | **Modest (n = 197)** |
| 1 – 2 | 211.1 (118.4) | 221.9 (105.2) | 174.3 (116.7) |
| 2 – 3 | 180.6 (110.6) | 196.8 (105.8) | 171.1 (101.1) |
| 3 – 4 | 190.9 (108.1) | 191.4 (105.6) | 165.2 (105.5) |
| 4 – 5 | 226.2 (134.4) | 215.5 (102.6) | 191.6 (104.9) |
| 5 – 6 | 219.8 (129.1) | 215.1 (101.7) | 187.7 (100.0) |
| 6 – 7 | 246.1 (141.4) | 216.2 (107.0) | 178.7 (90.40) |
| 7 – 8 | 319.3 (147.5) | 261.5 (122.0) | 229.6 (127.0) |
| 8 – 9 | 334.4 (144.6) | 297.3 (165.7) | 260.4 (145.4) |
| 9 – 10 | 355.0 (140.2) | 287.5 (145.8) | 252.3 (139.6) |
| 10 – 11 | 350.2 (191.4) | 331.6 (238.2) | 270.1 (147.2) |
| 11 – 12 | 422.3 (152.7) | 245.7 (162.0) | 263.0 (121.3) |
| 12 – 13* | - | 100 (0) | 284.7 (78.24) |

The results reported are the mean challenge test dose (standard deviation).

*At 12 – 13 years, there are no patients in the Striking group and only one patient in the Excellent group.
