## Supplementary Table 4 for "Dopa responsiveness in Parkinson’s disease"

***SUPPLEMENTARY TABLE 4***: Challenge test responses according to treatment response group and disease duration

| | | Disease duration in years | | | |
|---|---|---|---|---|---|
| | | **1 - < 4 years** | **4 - < 7 years** | **7- < 10 years** | **10-13** |
| **Striking** | **Number of patients, n (%)** | 26 (89.6%) | 26 (89.6%) | 19 (65.5%) | 6 (20.7%) |
| | **Challenge dose[a]** | 183.7 (101.7) | 219.4 (129.4) | 326.0 (146.1) | 382 (188.1) |
| | **Daily dose** | 503.2 (298.0) | 771.4 (435.1) | 1150 (424.1) | 1142 (534.8) |
| | **MDS UPDRS III on** | 9.1 (6.0) | 9.9 (5.6) | 12.9 (8.5) | 17.8 (21.2) |
| | **MDS UPDRS III off** | 20.7 (10.1) | 27.3 (13.5) | 28.8 (15.3) | 35.3 (31.8) |
| | **Percent response** | 56.3 (16.8) | 62.7 (11.3) | 57.1 (10.9) | 55.7 (17.8) |
| | **Off time present, n (%)** | 22 (37.9%) | 51 (78.5%) | 35 (94.6%) | 8 (100%) |
| | **Dyskinesia present, n (%)** | 12 (20.7%) | 40 (61.5%) | 24 (64.9%) | 6 (75%) |
| | **MDS UPDRS IV (total)** | 1.0 (1.3) | 2.6 (2.3) | 4.7 (2.8) | 6.4 (2.9) |
| | **MoCA (total)** | 27.7 (1.9) | 28.0 (1.7) | 28.3 (2.0) | 27.8 (2.5) |
| | **Number of tests, n** | 58 | 65 | 37 | 8 |
| | **Tests where response ≥24.5%** | 54 (93.1%) | 63 (96.9%) | 37 (100%) | 7 (87.5%) |
| **Excellent** | **Number of patients** | 70 (63.6%) | 89 (80.9%) | 81 (73.6%) | 31 (28.2%) |
| | **Challenge dose** | 186.9 (104.6) | 216.9 (105.8) | 274.7 (137.0) | 313.3 (229.1) |
| | **Daily dose** | 484.7 (317.1) | 673.4 (331.9) | 878.3 (415.4) | 952.6 (592.5) |
| | **MDS UPDRS III on** | 16.1 (6.8) | 16.3 (6.4) | 19.3 (8.9) | 21.7 (10.5) |
| | **MDS UPDRS III off** | 23.2 (9.1) | 28.1 (9.9) | 33.7 (12.2) | 39.0 (14.0) |
| | **Percent response** | 29.4 (13.0) | 42.2 (10.0) | 43.0 (13.9) | 45.2 (14.0) |
| | **Off time present, n (%)** | 50 (36.2%) | 150 (72.1%) | 126 (84%) | 29 (72.5%) |
| | **Dyskinesia present, n (%)** | 10 (7.2%) | 85 (40.9%) | 93 (62%) | 22 (55%) |
| | **MDS UPDRS IV (total)** | 1.0 (1.7) | 2.6 (2.4) | 5.0 (3.4) | 5.6 (4.8) |
| | **MoCA (total)** | 26.5 (2.8) | 26.7 (3.5) | 27.2 (3.3) | 28.0 (3.0) |
| | **Number of tests** | 138 | 208 | 150 | 40 |
| | **Tests where response ≥24.5%** | 93 (67.4%) | 181 (87%) | 124 (82.7%) | 36 (90%) |
| **Modest** | **Number of patients** | 113 (57.4%) | 141 (71.6%) | 131 (66.5%) | 64 (32.5%) |
| | **Challenge dose** | 157.2 (99.1) | 183.7 (103.8) | 232.5 (134.8) | 264.9 (139.0) |
| | **Daily dose** | 451.9 (298.7) | 626.3 (360.2) | 760.2 (395.8) | 968.9 (436.7) |
| | **MDS UPDRS III on** | 25.7 (9.6) | 29.1 (10.5) | 29.4 (11.0) | 29.8 (11.7) |

| | | | | | |
|---|---|---|---|---|---|
| | **MDS UPDRS III off** | 31.0 (11.2) | 35.3 (11.9) | 37.4 (12.5) | 41.8 (15.2) |
| | **Percent response** | 16.4 (13.4) | 17.3 (10.1) | 21.2 (13.4) | 27.9 (14.2) |
| | **Off time present, n (%)** | 39 (19.2%) | 184 (59.2%) | 160 (72.1%) | 53 (62.4%) |
| | **Dyskinesia present, n (%)** | 20 (9.9%) | 100 (32.2%) | 102 (45.9%) | 38 (44.7%) |
| | **MDS UPDRS IV (total)** | 0.7 (1.6) | 1.6 (2.0) | 2.8 (2.7) | 3.5 (3.2) |
| | **MoCA (total)** | 25.6 (2.8) | 25.5 (3.7) | 26.0 (3.6) | 26.0 (4.2) |
| | **Number of tests** | 203 | 311 | 222 | 85 |
| | **Tests where response ≥24.5%** | 60 (29.6%) | 84 (27%) | 95 (42.8%) | 49 (57.6%) |
| **P values (Differences across 3 clusters)** | **Challenge dose** | 0.05 | 0.03 | 0.001 | 0.26 |
| | **Daily dose** | 0.42 | 0.09 | 0.0001 | 0.47 |
| | **MDS UPDRS III on** | <0.0001 | <0.0001 | <0.0001 | 0.001 |
| | **MDS UPDRS III off** | <0.0001 | <0.0001 | 0.004 | 0.25 |
| | **Percent response** | <0.0001 | <0.0001 | <0.0001 | <0.0001 |
| | **Off time present, n (%)** | 0.05 | 0.04 | 0.001 | 0.12 |
| | **Dyskinesia present, n (%)** | 0.07 | 0.02 | 0.02 | 0.45 |
| | **MDS UPDRS IV (total)** | 0.03 | 0.008 | <0.0001 | 0.03 |
| | **MoCA (total)** | 0.0004 | 0.0001 | 0.00247 | 0.0243 |
| | **Tests where response ≥24.5%** | <0.0001 | <0.0001 | <0.0001 | 0.0003 |

MDS UPDRS: Movement Disorder Society Unified Parkinson's Disease Rating Scale.

[a]Values are mean (standard deviation) unless otherwise indicated.
